# Antibiotic use in the past 8 years and gut microbiota composition

**DOI:** 10.1101/2024.10.14.24315441

**Authors:** Gabriel Baldanzi, Anna Larsson, Sergi Sayols-Baixeras, Koen F. Dekkers, Ulf Hammar, Diem Nguyen, Tíscar Graells, Shafqat Ahmad, Camila Gazolla Volpiano, Guillaume Meric, Josef D. Järhult, Thomas Tängdén, Jonas F. Ludvigsson, Lars Lind, Johan Sundström, Karl Michaëlsson, Johan Ärnlöv, Beatrice Kennedy, Marju Orho-Melander, Tove Fall

## Abstract

**BACKGROUND:** Disruptions in gut microbiota have been implicated in cardiometabolic disorders and other health outcomes. Antibiotics are known gut microbiota disruptors, but their long-term consequences on taxonomic composition of the gut microbiome remain underexplored.

**Methods:** We investigated associations between register-based oral antibiotic use over 8 years and gut microbiota composition assessed with fecal shotgun metagenomics in 15,131 adults from the Swedish population-based studies SCAPIS, MOS, and SIMPLER. We applied multivariable regression models with the number of prescriptions in three pre-specified periods before fecal sampling (<1 year, 1–4, 4–8 years) as the main exposures and adjusted for sociodemographics, lifestyle, and comorbidities. Secondary analyses included participants with only one antibiotic course or none.

**Results:** Antibiotic use <1 year before fecal sampling was associated with the greatest reduction in gut microbiota species diversity; however, antibiotic use 1–4 years and 4–8 years earlier was also associated with decreased diversity. Clindamycin, fluoroquinolones, and flucloxacillin accounted for most of the associations between antibiotic use and the abundance of individual species across all periods. Use of these three antibiotics 4–8 years earlier was associated with altered abundance of 10–14% of the species studied; use of penicillin V, extended-spectrum penicillins, and nitrofurantoin were associated with altered abundance of only a few species. Similar results were found when comparing one antibiotic course 4–8 years before sampling vs. none in the past 8 years.

**CONCLUSION:** Commonly prescribed antibiotics like clindamycin, fluoroquinolones, and the narrow-spectrum flucloxacillin appear to have long-lasting consequences for the gut microbiota.

---

Recurrent and long-term use of antibiotics has been associated with an increased risk of overweight and obesity,^1,2^ type 2 diabetes,^1,3,4^ and cardiovascular disease^5^ in observational studies, potentially due to disruptions to the gut microbiota.^6^ This hypothesis is supported by evidence linking the gut microbiota to human health, including obesity,^7^ cardiometabolic disorders,^8,9^ and autoimmune conditions.^10^

Smaller intervention studies in healthy volunteers have reported drastic alterations in the gut microbiota a few days after a course of oral antibiotics, in particular reductions in species diversity^11^ and microbial gene richness.^12^ Other short-term alterations include increased abundance of potential pathogens such as *Escherichia coli*,^13^ reduced abundance of the genera *Dialister, Veillonella*, and *Eubacterium*,^14^ enrichment of antimicrobial-resistance genes,^11^ and increased risk of *Clostridium difficile* infection.^15^ Most of these alterations tend to reverse between 2 and 6 months after the antibiotic use,16 although more persistent alterations have been reported.^17^

Despite the frequent use of antibiotics, population-based investigations examining their long-term consequences on gut microbiota have not been conducted at scale.16 In this study, we assessed how antibiotic use over the 8 years before fecal sampling affected gut microbiota composition. Therefore, we combined information from the National Prescribed Drug Register (NDPR), which captures all antibiotics dispensed to outpatients in Sweden, with gut microbiota data obtained by fecal deep shotgun metagenomics in three Swedish population-based studies.

## METHODS

### STUDY POPULATION

The study population included participants from the population-based Swedish CArdioPulmonary bioImage Study (SCAPIS),^18^ Malmö Offspring Study (MOS),^19^ and Swedish Infrastructure for Medical Population-based Life-course and Environmental Research (SIMPLER).^20^ Fecal samples from 17,928 participants were collected between 2018 and 2012, sequenced, and passed quality control.^21^ Fecal samples were collected at home and kept in the freezer until the scheduled visit at the test center. Information on comorbidities was obtained through self-reporting, the NDPR, and the National Patient Register. Cohort-specific details are provided in the Supplemental Methods.

### ANTIBIOTIC EXPOSURE

All oral antibiotics dispensed to outpatients in Sweden require a prescription and are registered in the NDPR.^22^ We retrieved information on all dispensed prescriptions with the Anatomical Therapeutic Chemical code J01 (antibacterials for systemic use) and classified them as beta-lactamase-sensitive penicillins (J01CE), beta-lactamase-resistant penicillins (J01CF), extended-spectrum penicillins (J01CA), penicillins combined with beta-lactamase inhibitors (J01CR), cephalosporins (J01DB, J01DC, J01DD), macrolides (J01FA), tetracyclines (J01A), lincosamides (J01FF), fluoroquinolones (J01MA), sulfonamides and trimethoprim (J01E), and nitrofurantoin (J01XE01). In Sweden, amoxicillin with clavulanic acid was the only oral penicillin combination available, penicillin V was the only beta-lactamase-sensitive penicillin, flucloxacillin was the only beta-lactamase-resistant penicillin, and clindamycin was the only lincosamide. The only extended-spectrum penicillins were amoxicillin and pivmecillinam. Because the NDPR was initiated on July 1, 2005^22^ and recruitment in SCAPIS and MOS started in 2013, we limited the history of antibiotic use to the 8 years before the study site visit. Exposures were divided into three periods: <8 and ≥4 years, <4 years and ≥1 year, and <1 year before the study site visit.

### EXCLUSION CRITERIA

The exclusion criteria included a site visit before July 1, 2013 (incomplete history of antibiotic use in the past 8 years), an antibiotic prescription in the 14 days before the site visit, use of antibiotics to treat acne/rosacea or as prophylaxis for urinary tract infection at the time of fecal sampling, diagnosis of chronic pulmonary disease (i.e., chronic pulmonary obstructive disease, chronic bronchitis, and emphysema) or inflammatory bowel disease, as these conditions often entail a recurrent need for antibiotics and substantially alter the gut microbiota.^23,24^ Complete information on exclusion criteria is provided in the Supplemental Methods.

### FECAL METAGENOMICS

SCAPIS and MOS fecal samples were sent to Clinical Microbiomics A/S (Copenhagen, Denmark) for DNA extraction and shotgun metagenomic sequencing with Illumina NovaSeq6000 system (Illumina, USA).8,21 The SIMPLER samples were sent to the Centre for Translational Microbiome Research at the Karolinska Institute (Stockholm, Sweden) and sequenced with DNBSEQ G400 or T7 sequencing instrument (MGI Tech Co.). The metagenomic reads from all three studies were analyzed by Clinical Microbiomics using their Human Profiler (CHAMP)^25^ and delivered as relative abundance matrix of species taxonomically annotated with Genome Taxonomy Database (GTDB) database release 214.^26^ The relative abundance of species was subjected to centered log-ratio transformation after addition of a pseudo-value equal to the minimal non-zero value. In total, we analyzed 1,262 species with a relative abundance >0.01% in >1% of the participants in the three studies (see Supplemental Methods for details). Species diversity in each sample was assessed by three alpha diversity metrics: Shannon index, species richness, and inverse Simpson index. While richness represents the number of species, the Shannon and inverse Simpson metrics account for both the richness and evenness of the abundances.

### STATISTICAL ANALYSIS

Our assumptions about effects of temporally stable covariates on our exposure and outcomes were displayed on a directed acyclic graph (Fig. S1).27 Some potential time-varying covariates were primarily collected at the time of fecal sampling and thus after the antibiotic exposure (see Supplemental Methods for detailed covariate information). D-separation criteria were applied to the graph to select covariates to the basic model, which included age, sex, education, smoking, country of birth, and site-specific DNA extraction plates. The full model included conditions that may increase the risk of bacterial infection: body mass index, diagnosis of any type of diabetes, autoimmune rheumatologic diseases, and previous cancer at any site. Proton-pump inhibitor use within 1 year before fecal sampling was also included in the full model, given its strong association with the gut microbiota.^24^ For the multivariable regression models, microbiota species diversity metrics and the abundance of each of species were modeled individually as the dependent variable. The number of courses of each antibiotic class in the three periods (4–8 years, 1–4 years, <1 year) before fecal sample collection were included as independent variables in the same model:

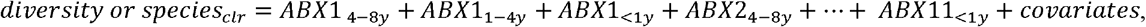

where ABX1–ABX11 are the number of courses of the 11 antibiotic classes, diversity is the gut microbiota species diversity, and species_clr_ is the centered log-ratio–transformed species abundance. The data from each cohort were analyzed separately and then subjected to inverse variance-weighted fixed-effects meta-analyses. Linear regression models were used for SCAPIS and SIMPLER; linear mixed-effects models with family as a random intercept were used for MOS. The generalized variance inflation factor was calculated to determine whether collinearity affected models. Multiple testing was accounted for using the Benjamini-Hochberg method, a false-discovery rate of 5% (q-value <0.05) was considered significant.^28^ In meta-analysis of results with a q-value <0.05 and Cohran’s Q p-value <0.05, we investigated the presence of influential observations (see Supplemental Methods). Additional analyses included estimated marginal mean diversity of gut microbiota species after each additional antibiotic course; association between antibiotics prescribed after fecal sampling (negative control) and species diversity, and associations in participants with 1 or no antibiotic prescription during the 8 years before sampling (see Supplemental Methods).

We performed two sensitivity analyses in SCAPIS and in SIMPLER using data available from the National Patient Register. In the first, participants who had been hospitalized for a condition likely to be treated with antibiotics (Table S1) were excluded. In the second, participants who had been hospitalized for any reason in the previous 8 years were excluded.

R version 4.3.2. was used for all analyses. Missing data were handled with complete-case analysis, as the amount of missingness was small and mostly random (Figure S2).

### ETHICS

Ethical approval was obtained from the Swedish Ethical Review Authority (DNR 2018-315 B and amendments 2020-06597 and 2022-06460-02, DNR 2012-594 and the amendment 2017-768 and 2020-05611, DNR 2022-06137-01 and amendment DNR 2023-04785-02). All participants from each of the three studies provided written informed consent.

## RESULTS

### STUDY POPULATION AND ANTIBIOTIC USE

After exclusions, the study population consisted of 8,576 participants from SCAPIS, 4,831 from SIMPLER, and 1,724 from MOS (Table 1 and Figure S2). The most prescribed antibiotics were penicillin V, extended-spectrum penicillins, and tetracyclines. The frequency of antibiotic use in the three studies was lower than in the general adult population in Sweden (Table 2).^29^ The proportion of participants who used any antibiotic at least once in the past 8 years varied from 70% in SCAPIS to 74% in SIMPLER. Correlation plots for the number of antibiotic courses in the three periods are shown in Figures S3–S5.

**Table 1.** Characteristics of Participants at the Time of Fecal Sampling.

| Characteristics | Number of Antibiotic Courses in Past 8 Years |  |  |  |  |  |
| --- | --- | --- | --- | --- | --- | --- |
|  | SCAPIS |  | SIMPLER |  | MOS |  |
|  | 0–1 | ≥2 | 0–1 | ≥2 | 0–1 | ≥2 |
| Basic model (N) | 4490 (52.4%) | 4086 (47.6%) | 2329 (48.2%) | 2502 (51.8%) | 854 (49.5%) | 870 (50.5%) |
| Age (years) | 57.1 [53.5;61.1] | 58.0 [54.0;61.6] | 72.0 [70.0;75.0] | 73.0 [71.0;75.0] | 39.5 [27.1;51.8] | 40.2 [27.9;52.4] |
| Female | 2044 (45.5%) | 2426 (59.4%) | 1008 (43.3%) | 1289 (51.5%) | 366 (42.9%) | 540 (62.1%) |
| Smoker |  |  |  |  |  |  |
| Never | 2504 (55.8%) | 1941 (47.5%) | 1229 (52.8%) | 1268 (50.7%) | 568 (66.5%) | 491 (56.4%) |
| Former | 1460 (32.5%) | 1576 (38.6%) | 932 (40.0%) | 1045 (41.8%) | 177 (20.7%) | 237 (27.2%) |
| Current | 526 (11.7%) | 569 (13.9%) | 168 (7.2%) | 189 (7.6%) | 109 (12.8%) | 142 (16.3%) |
| Highest education |  |  |  |  |  |  |
| Compulsory | 401 (8.9%) | 397 (9.7%) | 987 (42.4%) | 916 (36.6%) | 42 (4.9%) | 64 (7.4%) |
| Upper secondary | 1984 (44.2%) | 1835 (44.9%) | 681 (29.2%) | 772 (30.9%) | 483 (56.6%) | 481 (55.3%) |
| University | 2105 (46.9%) | 1854 (45.4%) | 661 (28.4%) | 814 (32.5%) | 329 (38.5%) | 325 (37.4%) |
| Born in Scandinavia* | 3773 (84.0%) | 3401 (83.2%) | 2244 (96.4%) | 2411 (96.4%) | 843 (98.7%) | 857 (98.5%) |
| Full model | 4395 (52.3%) | 4014 (47.7%) | 2327 (51.8%) | 2499 (49.3%) | 808 (49.3%) | 832 (50.7%) |
| BMI (kg/m<sup>2</sup>) | 26.3 [24.1;29.1] | 26.8 [24.1;29.9] | 25.7 [23.4;28.3] | 26.3 [23.9;29.0] | 25.0 [22.8;27.9] | 25.1 [22.4;28.6] |
| Diabetes | 330 (7.5%) | 362 (9.0%) | 151 (6.5%) | 219 (8.8%) | 19 (2.4%) | 35 (4.2%) |
| Autoimmune rheumatologic diseases | 107 (2.4%) | 203 (5.1%) | 32 (1.4%) | 61 (2.4%) | 16 (2.0%) | 38 (4.6%) |
| Previous cancer | 119 (2.7%) | 210 (5.2%) | 125 (5.4%) | 320 (12.8%) | 15 (1.9%) | 20 (2.4%) |
| PPI | 327 (7.4%) | 644 (16.0%) | 265 (11.4%) | 560 (22.4%) | 54 (6.7%) | 94 (11.3%) |

\*In MOS, “born in Sweden” was used instead.
Continuous variables are presented as median [25<sup>th</sup>–75<sup>th</sup> percentile], and categorical variables are presented as n (%).
SCAPIS: Swedish CARDioPulmonary bioImage Study; SIMPLER: Swedish Infrastructure for Medical Population-based Life-course Enviromental Research; MOS: Malmö Offspring Study; BMI: body mass index; PPI: proton-pump inhibitor use in the past year.

**Table 2.** Antibiotic Use in Population-Based Studies and in Sweden.

| **Antibiotic** | **Population-Based Studies** |  |  | **Sweden** |  |  |
| --- | --- | --- | --- | --- | --- | --- |
|  | **SCAPIS** | **SIMPLER** | **MOS** | **Age 20–49 y** | **Age 50–69 y** | **Age >70 y** |
| Any | 183.1 | 194.2 | 160.7 | 163.7 | 199.5 | 284.1 |
| Penicillin V | 69.4 | 59.8 | 65.0 | 65.0 | 68.0 | 69.4 |
| Flucloxacillin | 20.1 | 26.1 | 18.6 | 23.1 | 28.1 | 48.5 |
| Extended-spectrum penicillins | 37.9 | 44.9 | 29.0 | 30.9 | 43.9 | 88.4 |
| Amoxicillin/clavulanic acid | 3.8 | 2.3 | 1.2 | 3.8 | 5.0 | 6.2 |
| Cephalosporins | 5.9 | 4.1 | 9.9 | 5.2 | 4.4 | 8.7 |
| Macrolides | 5.0 | 2.5 | 8.7 | 6.7 | 6.1 | 5.2 |
| Clindamycin | 13.1 | 8.3 | 12.8 | 10.5 | 13.6 | 17.4 |
| Tetracyclines | 36.0 | 31.5 | 35.4 | 28.7 | 39.8 | 44.5 |
| Fluoroquinolones | 16.6 | 28.4 | 8.1 | 10.2 | 22.9 | 44.5 |
| Sulfamethoxazole-trimethoprim | 4.7 | 10.1 | 1.7 | 3.7 | 9.3 | 21.6 |
| Nitrofurantoin | 18.3 | 26.3 | 16.8 | 15.7 | 21.0 | 43.8 |

Number of individuals (per 1000) who used antibiotics in the year before fecal sampling and in the Swedish population in 2015. Study participants are those with complete data for covariates in the basic model.

### RECENT AND PAST USE OF ANTIBIOTICS WERE ASSOCIATED WITH A LOWER DIVERSITY OF GUT MICROBIOTA SPECIES

In general, effect estimates were consistent between the basic and the full model (Spearman correlation = 0.91, Table S2) and across cohorts (Figure S6). If not stated, we refer to full model results. The estimated diversity of gut microbiota species decreased with each additional antibiotic course within the three periods. The estimated decrease was greater for the first two courses than for the third and fourth courses (Figure 1A). Seven of the 11 antibiotic classes used <1 year previously were associated with a lower diversity in at least one metric (Figure 1B, Table S2). Clindamycin, fluoroquinolones, or flucloxacillin had the largest effect estimates. One course of clindamycin <1 year before fecal sampling was associated with an average of 52 fewer species detected (q-value=3.2×10^−23^). One course of fluoroquinolones or flucloxacillin was associated with an average of 22 and 20 fewer species detected (q-value=1.6×10^−8^ and 3.1×10^−7^, respectively). Fluoroquinolones, flucloxacillin, and tetracycline use 1–4 years and 4–8 years previously were also associated with lower diversity (Figure 1B), as was clindamycin and macrolide use 1–4 years, but not 4–8 years, previously. No associations were detected for or extended-spectrum penicillins (i.e., pivmecillinam and amoxicillin), amoxicillin/clavulanic acid, sulfamethoxazole-trimethoprim, and nitrofurantoin in any period (Table S2).

**Figure 1.**
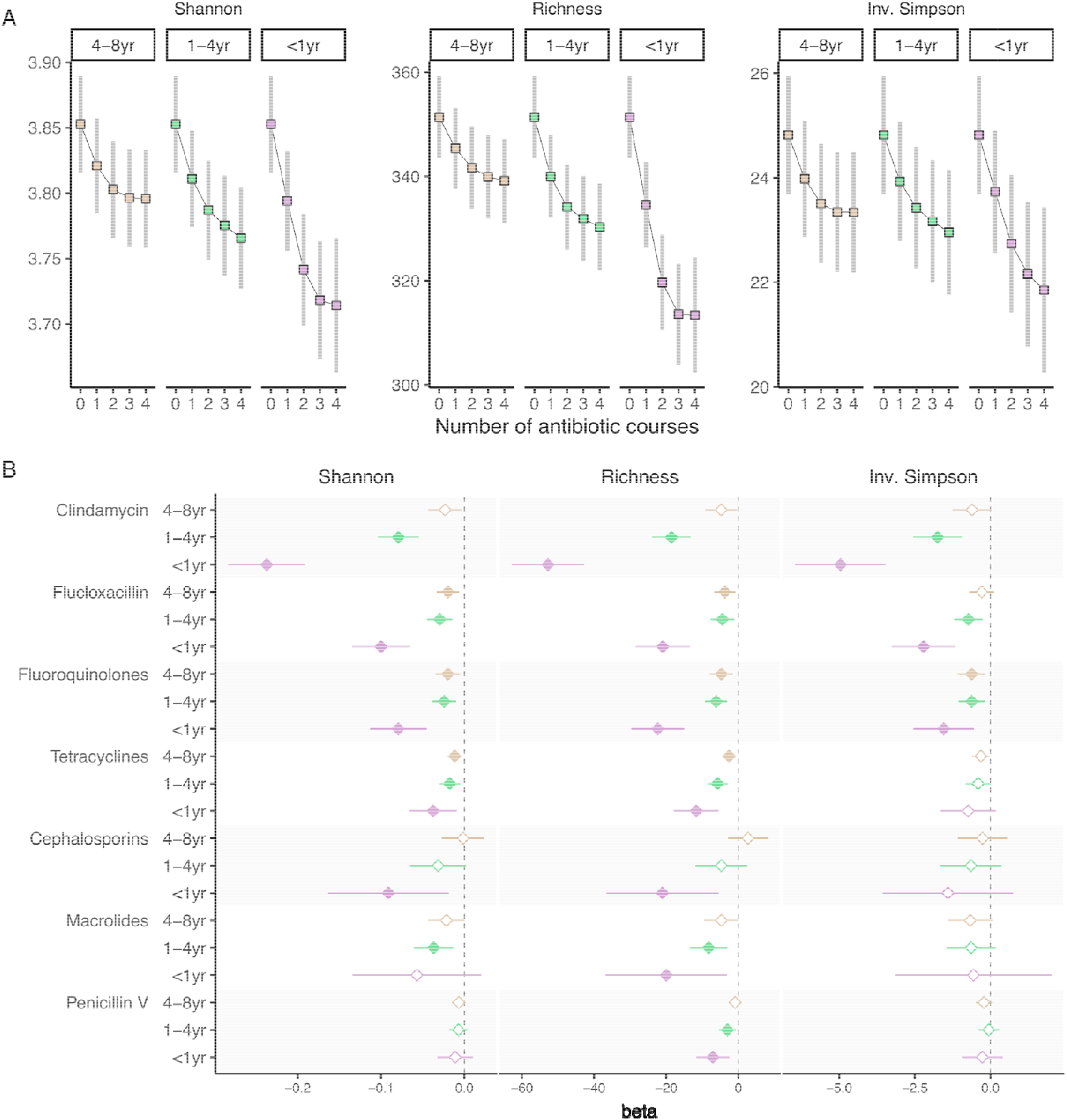
Antibiotic Use and Gut Microbiota Species Diversity. A. Microbiota diversity metrics (Shannon index, richness, and inverse Simpson index) for each additional course of any antibiotic within the periods 4–8 years, 1–4 years, and <1 year before fecal sampling. To estimate marginal means of the diversity metrics, the count of antibiotic courses was modeled with restricted cubic splines and adjusted for the full model. B. Associations between antibiotic use in the 8 years before fecal sampling and gut microbiota species diversity. Associations were investigated by using linear regression adjusted for age, sex, smoking, education, country of birth, site-specific analysis plate, body mass index, diabetes, previous cancer diagnosis, autoimmune rheumatologic diseases, and proton-pump inhibitor use. In MOS, linear mixed models were used with family as a random effect. Fixed-effects models were used for meta-analyses. Filled shapes indicate significant associations (5% false-discovery rate). Antibiotics with at least one such association are shown.

To assess whether the covariate adjustment controlled for the differences between exposed and nonexposed individuals, antibiotic use in the 1 year after fecal sampling was used as a negative control exposure. Antibiotic use after sampling was not associated with species diversity, indicating that the full model likely controlled for the most important confounding effects (Fig. S7 and Tables S3 and S4). No multicollinearity issues were detected (Table S2). In the sensitivity analyses, removing 550 individuals who were hospitalized for infection or 5,147 hospitalized for any reason in the past 8 years, the overall effect estimates were similar (Table S6 and Fig. S8). In an additional analysis, we restricted the sample to 7672 participants who had only one antibiotic course or none in the 8 years before fecal sampling, as this was a more homogenous population. A single course of clindamycin, cephalosporins, macrolides, fluoroquinolones, or flucloxacillin <4 years or 4–8 years before sampling was associated with lower microbiota species diversity (Fig. S9 and Table S6).

### CLINDAMYCIN, FLUCLOXACILLIN, AND FLUOROQUINOLONES HAD THE HIGHEST NUMBER OF ASSOCIATIONS WITH THE ABUNDANCE OF GUT MICROBIOTA SPECIES

Clindamycin, flucloxacillin, and fluoroquinolones accounted for most of the associations between antibiotic use and individual species abundances (Fig. 2 and Table S7). We discarded 37 associations driven by single influential observations (i.e.; p-value >0.05 after exclusion of one individual from each cohort). Regression coefficients of the basic and full models were highly consistent (Spearman correlation = 0.97), hence, we refer to the results of the full model. Although the associations for antibiotic use <1 year before fecal sampling were the strongest in terms of effect estimates and p-values, many associations were also observed for antibiotic use 1–4 and 4–8 years before sampling (Figure 2A and Table S7). Clindamycin use <1 year before sampling was associated with 326 of the 1,262 species analyzed, flucloxacillin with 189 species, and fluoroquinolones with 184 species. For comparison, penicillin V, the most prescribed antibiotic, was associated with only 36 species. Most associations were in the negative direction (i.e., decreased abundance of the species), but positive associations were also observed. For example, clindamycin use 1–4 years before sampling was associated with reduced abundance of 226 and increased abundance of 111 species.

**Figure 2.**
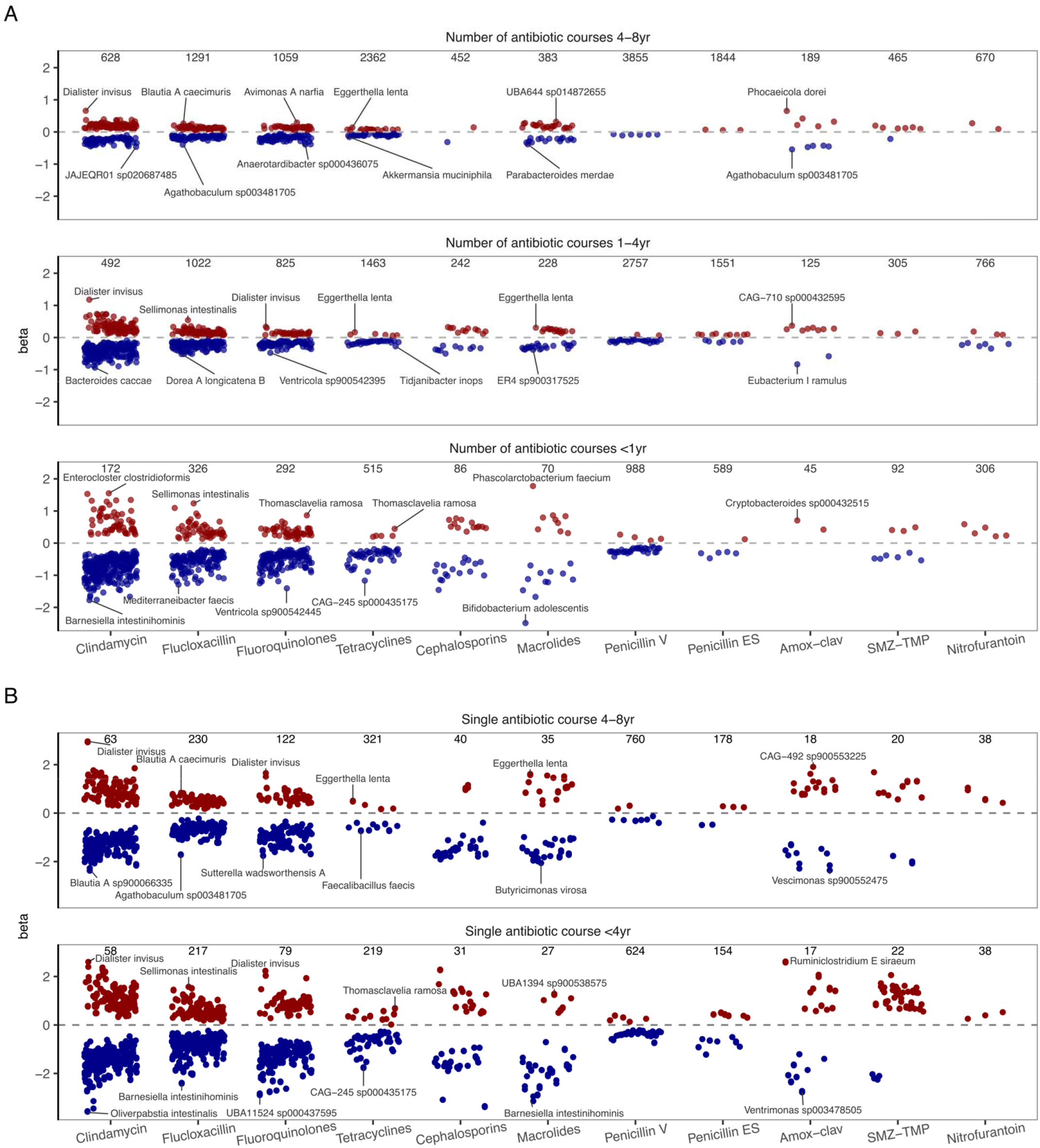
Antibiotic Use and Gut Microbiota Species Abundance. Associations were investigated by using linear regression adjusted for age, sex, smoking, education, country of birth, site-specific analysis plate, body mass index, diabetes, previous cancer diagnosis, autoimmune rheumatologic diseases, and proton-pump inhibitor use. In MOS, linear mixed models were used with family as a random intercept. Analyses were done separately by study; fixed-effects models were used for meta-analyses. Relative species abundances were subjected to centered log-ratio transformation. Only significant associations at a 5% false discovery rate are shown. Red circles indicate positive associations (increased species abundance); blue circles indicate negative associations (decreased species abundance). A. The number of prescriptions in the three periods for each antibiotic class (4–8 years, 1–4 years, <1 year before fecal sampling) were included in the same model. The numbers above the circles indicate number of participants who used the antibiotic at least once in that period. B. Analysis of participants who had only one or no antibiotic course 4–8 years or <4 years before fecal sampling. The numbers above the circles indicate numbers of participants who used the antibiotics in that period. Penicillin ES: penicillins with extended spectrum; Amox-clav: amoxicillin/clavulanic acid; SMZ-TMP: sulfamethoxazole-trimethoprim.

Flucloxacillin use was mostly associated with alterations in bacteria in the phylum *Bacillota* A, orders *Lachnospirales* and *Oscillopirales*, which are mostly Gram-positive bacteria. In contrast, clindamycin and especially fluoroquinolones were associated with reduced abundance of bacteria from a more diverse range of phyla and orders (Figure 3). Clindamycin, flucloxacillin, and fluoroquinolones continued to account for most of the significant associations after exclusion of individuals who had been hospitalized for infection or for any reason in the past 8 years (Fig. S10 and Table S8).

**Figure 3.**
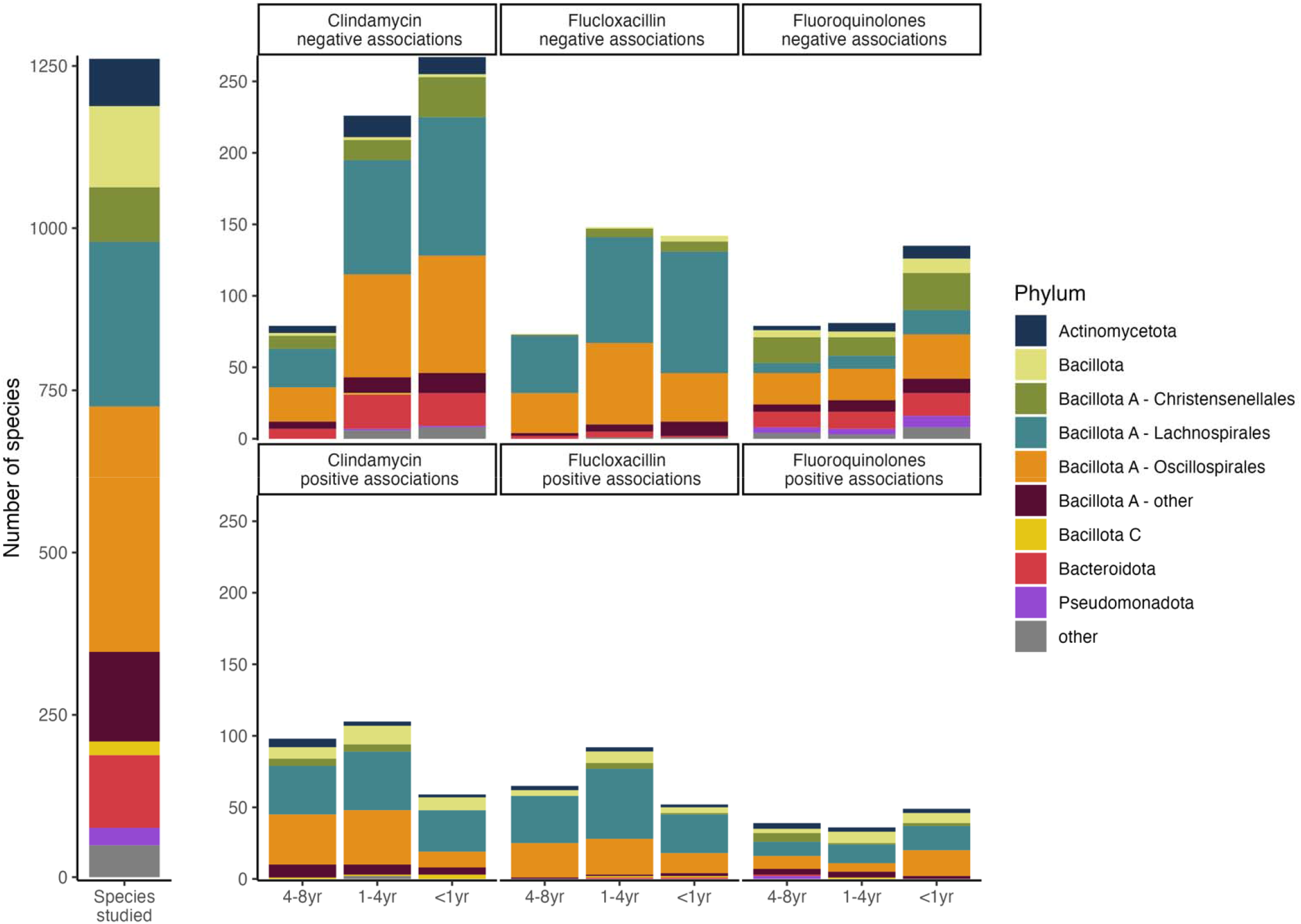
Taxonomy of Species Significantly Associated with Clindamycin, Flucloxacillin, and Fluoroquinolones in the Full Model at a 5% False Discovery Rate. The species studied are the 1,262 species that were present in at least 1% of the participants at a relative abundance >0.01%. The *Bacillota* A phylum was subdivided into the main orders.

In the analysis restricted to individuals who had only one or no antibiotic course in the previous 8 years, clindamycin, flucloxacillin, and fluoroquinolones were again the antibiotics most associated with species abundance (Figure 2B and Table S9). A single course of clindamycin, flucloxacillin, or fluoroquinolones <4 years before sampling was associated with 234, 286, and 170 species, respectively. A single course of these antibiotics 4–8 years before sampling was associated with 190, 147, and 91 species, respectively.

## DISCUSSION

In this population-based study of 15,131 participants, we investigated the association between antibiotic use in the 8 years before fecal sampling and the composition of the gut microbiota. Three findings stand out. First, while the strongest associations were found for antibiotics used <1 year before sampling, antibiotics used 1–4 years and 4–8 years before were also associated with lower diversity and altered abundance of species. Second, the associations identified with species abundance were mainly related to three antibiotics—clindamycin, flucloxacillin, and fluoroquinolones. Third, even a single course of antibiotics 4–8 years before sampling was associated with alterations in the gut microbiota. These findings support the notion that the effect of antibiotics on the gut microbiota may persist for several years.

Alterations of the gut microbiota after antibiotic use have been characterized by loss and lower diversity of species, ^30,31^ altered metabolic functions,^31^ reduced colonization resistance against pathogenic species such as *C. difficile*,^32^ and blooming of antibiotic-resistant clones.^33^ Lower species diversity has been associated with a range of traits and diagnoses, such as obesity, diabetes, and inflammatory bowel disease.^31,33,34^ The altered microbiota might also promote systemic inflammation by impairing the gut barrier^35^ and reducing the mucus layer.^36^ Altogether, antibiotic-induced alterations in gut microbiota may contribute to the development of diseases. Notably, studies have shown connections between antibiotic use and obesity, diabetes,^1,3,4^ and cardiovascular disease.^5^ We found, for instance, that use of clindamycin, fluoroquinolones, and flucloxacillin was associated with a greater abundance of *Enterocloster bolteae, E. citroniae* (previously *Clostridium bolteae*, and *C. citroniae*), *Flavonifractor plautii, Ruminococcus gnavus, Eggerthella lenta*, and *Collinsela stercoris*. These species have been associated with a higher body mass index, type 2 diabetes, and higher serum triglycerides.^37–39^

How well the microbiota recovers after antibiotics is not fully understood. A partial recovery often occurs within weeks^11,16,40^ but might take years.^17,41,42^ In intervention studies (n = 6–66), the duration of the reduction in the species diversity varied by antibiotic class.^11,13,40,43^ In one study, diversity remained reduced for 4 months after clindamycin use and 12 months after fluoroquinolone use, but no effect was observed after amoxicillin, an extended-spectrum penicillin.^44^ Similarly, clindamycin and fluoroquinolone use was associated with lower microbiota species diversity in the current study, but use of extended-spectrum penicillins was not.

Observational studies have described associations between a years-long history of antibiotic use and gut microbiota. In the MetaCardis consortium (n=2,173), antibiotic use in the past 5 years was ranked third in explaining variability in microbiota taxonomy, after diet and country of residence.^45^ In the Estonian Microbiome Cohort (n=2,509), the number of antibiotic courses in the last 10 years was associated with lower microbiota species diversity.^46^ However, neither study differentiated recent antibiotic use from past use or analyzed by antibiotic class. In our study, both recent use and antibiotic use 4–8 years before sampling were associated with the gut microbiota, and the associations differed by antibiotic classes.

The strong association between clindamycin use and gut microbiota could be explained by predominantly biliary excretion of this antibiotic and its broad effect on anaerobes.^47^ Fluoroquinolones are broad-spectrum antibiotics that especially target Gram-negative bacteria^48^ and their large impact on gut microbiota has been highlighted.^31^ Fluoroquinolones have a bioavailability above 70% and are mainly excreted via the kidneys.^49^ Additionally, repetitive use of fluoroquinolones is associated with incident diabetes.^3^ Both fluoroquinolones and clindamycin use are associated with a higher risk of *C. difficile* infection.^15^

Flucloxacillin, a beta-lactamase-resistant penicillin, has 50–70% bioavailability and is excreted mainly via the kidneys.^50^ Despite its narrow spectrum against Gram-positive bacteria, we found that flucloxacillin was strongly associated with the gut microbiota. Notably, most of the species associated with flucloxacillin use belonged to the *Bacillota* A phylum, which consists primarily of Gram-positive bacteria.

The NDPR was used to ascertain antibiotic use. This registry captures all antibiotics dispensed in Sweden to outpatients. Because antibiotics provided during hospitalization are not captured by the registry, underestimation of antibiotic use could have affected our results. However, sensitivity analyses after exclusion of participants hospitalized in the 8 years before sampling provided results similar to those of the full model. Another limitation is that the NDPR does not include treatment indications, which hampered analyses to distinguish the effect of antibiotics from the effects of infections. However, since antibiotics are mainly prescribed for airway, urinary tract, skin, and soft tissue infections,^51^ the possible effect of these infections on the gut microbiota is likely less pronounced than the effect of the antibiotics.

The main strengths of this study are the large study sample and the fecal metagenomic data from three population-based studies profiled using the same method, which allowed harmonization of species annotations and meta-analysis. A limitation of the data is that the exact date of fecal sample collection was not systematically recorded, so instead we used the date when the participant visited the test center. However, this should not have affected the long-term associations identified. In addition, since antibiotic use in Sweden is more restrictive than in many other countries, our findings may not directly apply to other populations. Certain antibiotics, such as combinations of penicillins with beta-lactamase inhibitors, are rarely prescribed in Sweden. Thus, we had limited statistical power for these antibiotics. The abundance of microbiota species was measured as relative abundances, the data type most commonly available in large population-based studies. Relative abundance might not reflect changes in the absolute abundance.^52^

In conclusion, we found evidence that certain antibiotics classes influence the gut microbiota for more than 4 years. Clindamycin, fluoroquinolones, and flucloxacillin were the classes that had the largest effects. Our results may inform future guidelines on antibiotic use, which should, when possible, prioritize antibiotics that have a lower impact on the gut microbiota.

## Data Availability

The data supporting the conclusions of this article were provided by the SCAPIS, SIMPER, and MOS and are not shared publicly due to confidentiality. Data will be shared upon reasonable request to the corresponding author only after permission from the Swedish Ethical Review Authority (https://etikprovningsmyndigheten.se) and from the boards of SCAPIS, (https://www.scapis.org/data-access), SIMPER (https://www.simpler4health.se), and MOS (https://www.malmo-kohorter.lu.se/malmo-offspring-study-mos). The code used for the statistical analysis and .csv files of the supplementary tables will be available at https://github.com/MolEpicUU/antibiot_gut. 

## Contributors

J.S., L.L, J.Ä., M.O-M., and T.F. obtained the funding for the study. G.B., S.S.-B., K.F.D., U.H., B.K., and T.F. planned and designed the study. G.B. performed the statistical analyses with support from S.S.-B., K.F.D., U.H., and T.F. G.B. wrote the first draft of the manuscript with contributions from A.L., M.O-M., and T.F. All authors helped interpret the results and revise the manuscript.

## Acknowledgments

Financial support was obtained in the form of grants from the European Research Council [ERC-STG-2018-801965 (T.F.); ERC-CoG-2014-649021 (M.O.-M.); ERC-STG-2015-679242 (J.G.S.)], the Swedish Heart-Lung Foundation [Hjärt-Lungfonden, 2019-0505 (T.F.); 2018-0343 (J.Ä.); 2020-0711 (M.O.-M.)], the Swedish Research Council [VR, 2019-01471 (T.F.), 2018-02784 (MO-M), 2018-02837 (M.O.-M.), 2019-01015 (J.Ä.), 2020-00243 (J.Ä.), 2022-01460 (S.A.), and EXODIAB 2009-1039 (M.O.-M.)], the Swedish Research Council for Sustainable Development [FORMAS, 2020-00989 (S.A.)], Göran Gustafsson foundation [2016 (T.F.)], Axel and Signe Lagerman’s foundation (T.F.), the A.L.F. governmental grant 2018-0148 (M.O.-M.), The Novo Nordic Foundation NNF20OC0063886 (M.O.-M.), The Swedish Diabetes Foundation DIA 2018-375 (M.O.-M.), Center of Clinical Research (CKF) in Region Dalarna (J.Ä.), Epihealth (S.A.), and governmental funding of clinical research within the Swedish National Health Service (J.G.S.). B.K. is supported by a Gullstrand fellow grant from the Uppsala University Hospital. We acknowledge the Swedish Heart-Lung Foundation, the main funding body of SCAPIS. Funding for the SCAPIS study was also provided by the Knut and Alice Wallenberg Foundation, the Swedish Research Council and VINNOVA (Sweden’s innovation agency), the University of Gothenburg and Sahlgrenska University Hospital, Karolinska Institutet and Stockholm County Council, Linköping University and University Hospital, Lund University and Skåne University Hospital, Umeå University and University Hospital, Uppsala University and University Hospital. We thank SIMPLER for the providing facilities and experimental support and Anna-Karin Kolseth and Niclas Håkansson for assistance. SIMPLER receives funding through the Swedish Research Council under grant no 2017-00644, 2017-06100, 2021-00160, and Stiftelsen Olle Engkvist Byggmästare. The computations and data handling were enabled by resources in project sens2019512 and simp2023007 provided by the National Academic Infrastructure for Supercomputing in Sweden (NAISS) at Uppsala Multidisciplinary Center for Advanced Computational Science (UPPMAX), funded by the Swedish Research Council through grant agreement no. 2022-06725

