## Supplemental Material for "Antibiotic use in the past 8 years and gut microbiota composition"

**Supplementary Appendix**

### Supplemental Methods

#### SCAPIS

The Swedish CArdioPulmonary bioImage Study (SCAPIS) enrolled 30,154 women and men aged 50–65 invited from a random sample of residents in areas adjacent to 6 academic hospitals in Sweden between 2013 and 2018.[1] Participation in SCAPIS entailed two visits to the test site. In the Uppsala and Malmö sites, participants were provided with material and instructions to collect a fecal sample at home close to the second visit, store it in the home freezer, and bring the sample at the second visit The median interval between visits was 9 days. If a participant failed to provide a fecal sample at the second visit, they were allowed to deliver it at a later time. The date of sampling was not consistently recorded. In total, 9,159 fecal samples were delivered to the research center by the second visit, 244 were delivered ≤7 days after the second visit, and additional 165 samples were delivered >7 days after the second visit. The delivery date was missing for 248 samples. Participants also answered an extensive questionnaire on lifestyle, diet, and health history. Blood samples were collected, and anthropometric measurements were conducted on-site. Proton-pump inhibitor use was based on the Prescribed Drug Register as a prescription with Anatomical Therapeutical Chemical (ATC) code A02BC in the year before fecal sampling. Proton-pump inhibitors are also sold without a prescription in Sweden; however, in smaller packages and for a higher price. Therefore, it can be assumed that most long-term proton-pump users have a prescription. Besides self-reported doctor diagnoses, the diagnoses of chronic obstructive pulmonary disease, chronic bronchitis, or emphysema were attributed to participants with the ICD-10 codes J41, J42, J43, or J44 in the National Patient Register in the 8 years before fecal sampling. Autoimmune rheumatologic disease was self-reported as having a diagnosis of rheumatoid arthritis, psoriatic arthritis, systemic lupus erythematosus, Sjögren's syndrome, or other inflammatory rheumatoid disease.

#### MOS

The Malmö Offspring Study (MOS)[2] includes the adult children and grandchildren of the population-based Malmö Diet and Cancer-Cardiovascular Cohort participants.[3] The recruitment occurred between 2013 and 2021. After the exclusion of 39 participants who were also part of SCAPIS, the present study included 2,223 participants enrolled until April 2017 whose fecal samples were analyzed with shotgun metagenomics and passed quality control. Information on comorbidities was self-reported. As in SCAPIS, participants in MOS were instructed to collect the fecal samples at home with the material provided and bring the sample to the study site on the second visit. Participants who did not provide a fecal sample at the second visit could provide a sample at a later time. Also in MOS, the date of sampling was not consistently recorded. In total, 1,573 samples were delivered by the second visit, 73 were delivered in the 7 days following the second visit, 10 were delivered >7 days after the second visit, and the delivery date was missing for 567 samples. Lifestyle, diet, health history, and comorbidities were assessed using questionnaires. Autoimmune rheumatologic diseases were self-reported as having a diagnosis of rheumatoid arthritis, psoriatic arthritis, systemic lupus erythematosus, Sjögren's syndrome, or other inflammatory rheumatoid disease. Anthropometric measurements and blood sample collection were performed during the study site visit. Use of proton-pump inhibitors was defined as a prescription starting with the ATC A02BC in the last year or self-reported use in the last 6 months.

**SIMPLER**

The Swedish Mammography Cohort (SMC), initiated in 1987, and the Cohort of Swedish Men (COSM), initiated in 1997, are two prospective cohorts in central Sweden that compose the Swedish Infrastructure for Medical Population-based Life-course and Environmental Research (SIMPLER).[4] In total, 2,843 women and 3,046 men provided fecal samples between 2012 and 2018. As fecal samples were analyzed together and the phenotype data have been harmonized by SIMPLER, we pooled data from the two cohorts. Participants received via mail the material and instructions to collect a fecal sample at home close to the date of the health examination on site. At the time of fecal sampling, participants answered a questionnaire that included questions on smoking, physical activity, and a food frequency questionnaire. In cases when smoking was not reported, the most recent smoking information was used, which could be the questionnaire in 2009 or 2019 depending on the date of fecal sampling. At the health examination, anthropometric measurements were performed. The diagnoses of chronic pulmonary disease (ICD-10 codes J41, J42, J43, or J44), inflammatory bowel disease (K50 or K51), autoimmune rheumatologic diseases (L405, M05, M06.0, M06.2, M06.3, M06.8, M06.9, M07.0, M07.1, M07.2, M07.3, M32.1, M32.8, M32.9, M35.0, and M33), and cancer (all C codes) were obtained using the National Patient Register. Diabetes diagnosis was defined as having a prescription for a medication starting with the ATC A10A (insulins and analogues) or A10B (blood glucose lowering drugs, excluding insulins) in the 18 months before fecal sampling. The use of proton-pump inhibitors was defined as a prescription with ATC code A02BC in the last year.

**Fecal metagenomics**

SCAPIS and MOS fecal samples were sent to Clinical Microbiomics A/S (Copenhagen, Denmark) for DNA extraction and shotgun metagenomic sequencing.[5,6] DNA extraction was performed using NucleoSpin® 96 Soil kits. Each round of extraction contained a negative and a positive control. Following DNA fragmentation and library preparation, sequencing was conducted with the Illumina NovaSeq6000 system (Illumina, USA). The average sequence depth was 25.3 million read-pairs for SCAPIS Uppsala samples and 26.3 million read-pairs for SCAPIS Malmö and MOS samples.

The fecal samples from SIMPLER were sent to the Centre for Translational Microbiome Research at the Karolinska Institute in Stockholm, Sweden. Before shipping, the samples were aliquoted into FluidX tubes containing 800μl of DNA/RNA Shield buffer (R1100-250, Zymo Research). The DNA extraction was conducted with the MagPure Stool kit (Magen Biotechnology Co.) and included a bead-beating step in a FastPrep-96™ at 1600 rpm for one minute. One negative and one positive control were added to each batch. Library preparation was performed using the MGIEasy FS DNA Library Prep Set kit. The libraries were sequenced in MGI Tech Co (Latvia) using DNBseq 2x100 bp paired-end sequencing on the DNBSEQ G400 or T7 sequencing instrument (MGI Tech Co.). The average sequence depth was 51 million read-pairs.

Clinical Microbiomics profiled the metagenomic sequences from SCAPIS, SIMPLER, and MOS using their Human Microbiome Reference HMR05 gene catalog,[7] which was mainly derived from prokaryotic metagenome-assembled genomes (MAGs) from the Unified Human Gastrointestinal Genome collection,[8] the Early-Life Gut Genomes catalog,[9] as well as selected genomes from NCBI and PATRIC. Eukaryotic species were identified from different sources, including a public list of pathogens and gut fungal species from the Human Microbiome Project.[10] The MAGs were taxonomically annotated with Genome Taxonomy Database (GTDB) database release 214.[11]

Metagenomic reads were analyzed using Clinical Microbiomics Human Profiler (CHAMP$^{TM}$).[7] Briefly, reads that mapped to the human reference genome GRCh38.p14 were removed using Bowtie2 (v 2.4.2). The non-host reads were mapped to the HMR05 gene catalog using BWA mem (v. 0.7.17). The relative abundance of each species (MAGs) was calculated based on the species signature genes with observed read counts within the expected 99% quantile and normalized sample-wise so that the total abundance of all species summed to 100%. The expected 99% quantile of read counts was calculated for each gene based on a negative binomial distribution with a mean proportional to the effective gene length and dispersion as log2(effective gene length). The species relative abundance was centered-log ratio (clr) transformed after adding a study-specific pseudo-value equal to the minimal non-zero abundance value. After transformation, the values equal to zero before transformation were replaced with the minimal non-zero transformed value for each species to avoid that species with an initial value equal to zero had different values after the transformation. We kept for subsequent analysis the 1,262 species with a relative abundance >0.01% in >1% of the participants in the three studies. A rarefied species abundance table was produced by random sampling, without replacement, of 190,977 gene counts per sample in SCAPIS and MOS and 641,964 gene counts in SIMPLER. The diversity of microbiota species in each sample was assessed using the rarefied table to calculate three alpha diversity metrics: Shannon index, species richness, and inverse Simpson index.

**Exclusion criteria**

In SCAPIS, 25 participants who did not consent to the data linkage with registry data were excluded. Participants with a site visit before July 1, 2013, were excluded as their history of antibiotic use in the past 8 years was not available. To exclude samples collected during antibiotic treatment, we excluded individuals with an antibiotic prescription in the 14 days preceding the study site visit, and participants with prescriptions patterns of antibiotic use for urinary tract infection prophylaxis or acne/rosacea treatment at the time of fecal sampling. Long-term medications are typically dispensed for three-month periods. Therefore, we excluded all participants with a dispensed methenamine prescription in the three months before the fecal sampling. Likewise, we excluded participants with dispensed nitrofurantoin or trimethoprim prescriptions in the 12 weeks before fecal sampling summing to at least 22.5 defined daily doses (DDD), which is equivalent to 50 mg and 100 mg, respectively, once a day for 12 weeks. To exclude long-term users of doxycycline for rosacea, we excluded all participants with a prescription of 40mg of doxycycline tablets in the 12 weeks before the fecal sampling, all participants with one or more prescriptions of 100mg of doxycycline tablets adding up to at least 84 DDD in the last 12 weeks (equivalent to 100mg/day for 12 weeks), at least 56 DDD in the last 8 weeks, or at least 42 DDD in last 6 weeks. To exclude long-term users of tetracycline or lymecycline for rosacea, we excluded all participants with dispensed prescriptions adding up to at least 42 DDD in the last 12 weeks (equivalent to 500mg/day or 300mg/day, respectively, for 12 weeks), at least 28 DDD in the last 8 weeks, or at least 21 DDD in the last 6 weeks.

Because the visit dates were available but not the exact date of fecal sampling, we excluded SCAPIS and MOS participants who a) had an antibiotic prescription between the two visits, b) provided fecal samples >7 days after the second visit, or c) had an interval of >60 days between visits given the uncertainty about sample collection date. Additionally, we excluded those with a diagnosis of chronic pulmonary disease (i.e., chronic pulmonary obstructive disease, chronic bronchitis, and emphysema), and/or inflammatory bowel disease (ulcerative colitis or Crohn’s disease), as these conditions often entail a recurrent need for antibiotics and have been associated with substantial alterations in the gut microbiota.[12,13]

**Covariate management**

Self-reported smoking status was grouped into never, former, or current. The highest education level achieved was categorized as compulsory, upper secondary, and university education. In SCAPIS and SIMPLER, the country of birth was grouped in Scandinavia, non-Scandinavian Europe, Asia, and others. In MOS, the country of birth was grouped into Sweden or another. Cancer diagnoses were categorized into no diagnosis, diagnosis in the same year or the year before, 2 to 4 years, and 5 to 8 years before the fecal sampling.

**Additional statistical analyses**

In meta-analysis results with a q-value <0.05 and Cohran's Q p-value <0.05, we investigated the presence of an influential observation in the individual studies. To identify the influential observations, after re-running the linear regression within each study, we calculated dfbetas for the antibiotic exposure with a q-value <0.05. Next, we re-ran the model after removing the observations with the highest dfbeta in each of the studies and meta-analyzed the estimates. If the new meta-analysis results had a p-value >0.05, the initial significant association was discarded.

To estimate the marginal mean gut microbiota species diversity associated with each additional antibiotic course within each period, we counted the number of prescriptions for all antibiotics in each period and modeled them as independent variables using restricted cubic splines with three knots and adjusted for the full model covariates.

To assess whether our full model sufficiently controlled for confounders, we used as a negative control exposure the association between antibiotic use in the year after fecal sampling and the gut microbiota species diversity, either adjusting for antibiotic use before the sampling or restricting to participants with no antibiotic before the sampling. We excluded 1,458 participants in SCAPIS for whom register data were unavailable for the year after fecal sampling.

Because the Prescribed Drug Register does not capture antibiotics prescribed to inpatients, we performed two sensitivity analyses in SCAPIS and in SIMPLER, where data on hospitalizations from the National Patient Register were available: 1) exclusion of participants who had been hospitalized in the 8 years before the fecal sampling due to a condition likely to be treated with antibiotics (Table S1), and 2) exclusion of participants who had been hospitalized due to any reason in the 8 years before fecal sampling.

Additionally, we investigated the association between a single antibiotic course and the gut microbiota by restricting our analysis to individuals with only one antibiotic prescription or none during the 8 years before fecal sampling. To ensure sufficient statistical power for antibiotic classes less often prescribed, the periods 1–4 and <1 year were merged in this analysis.

#### Supplemental Figures

**Figure S1. Directed Acyclic Graph**

Directed acyclic graph of the hypothetical causal diagram of the effect of antibiotic use on gut microbiota.

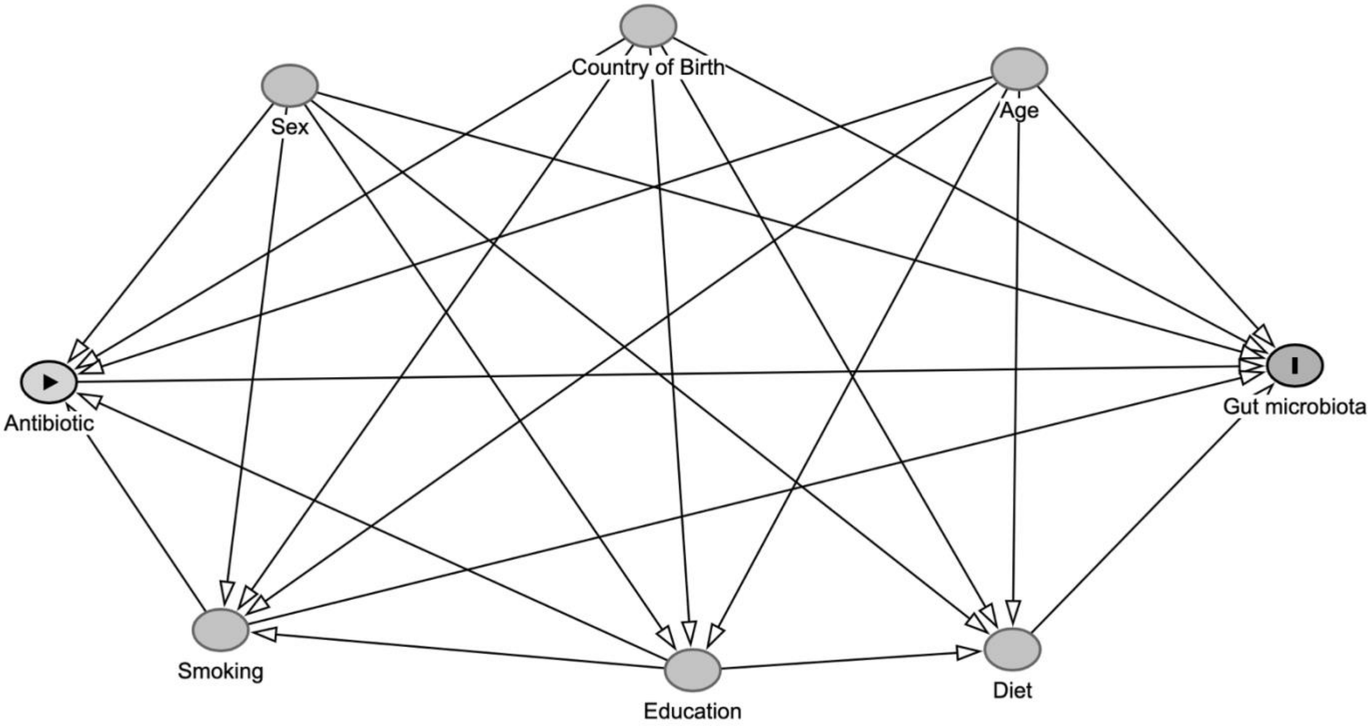

**Figure S2. Flowchart of Exclusion of Participants**

Flowchart of exclusion of participants and final sample size for the basic and full models. COPD: chronic pulmonary obstructive disease, IBD: inflammatory bowel disease.

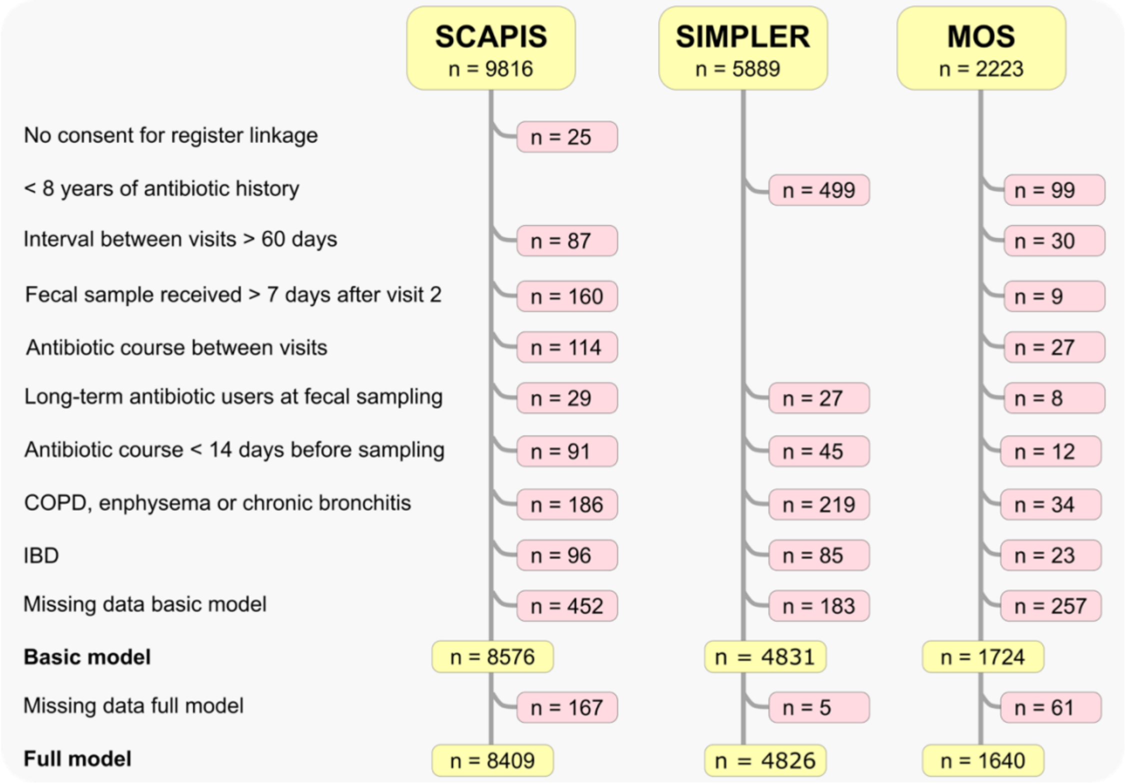

**Figure S3. Spearman Correlation in SCAPIS Between Antibiotic Classes**

Spearman correlation in SCAPIS between the number of prescriptions of each antibiotic class in the periods 4–8 years, 1–4 years, <1 year before the fecal sampling. Penicillin ES: extended-spectrum penicillins; Amox-clav: amoxicillin/clavulanic acid; SMZ-TMP: sulfamethoxazole-trimethoprim.

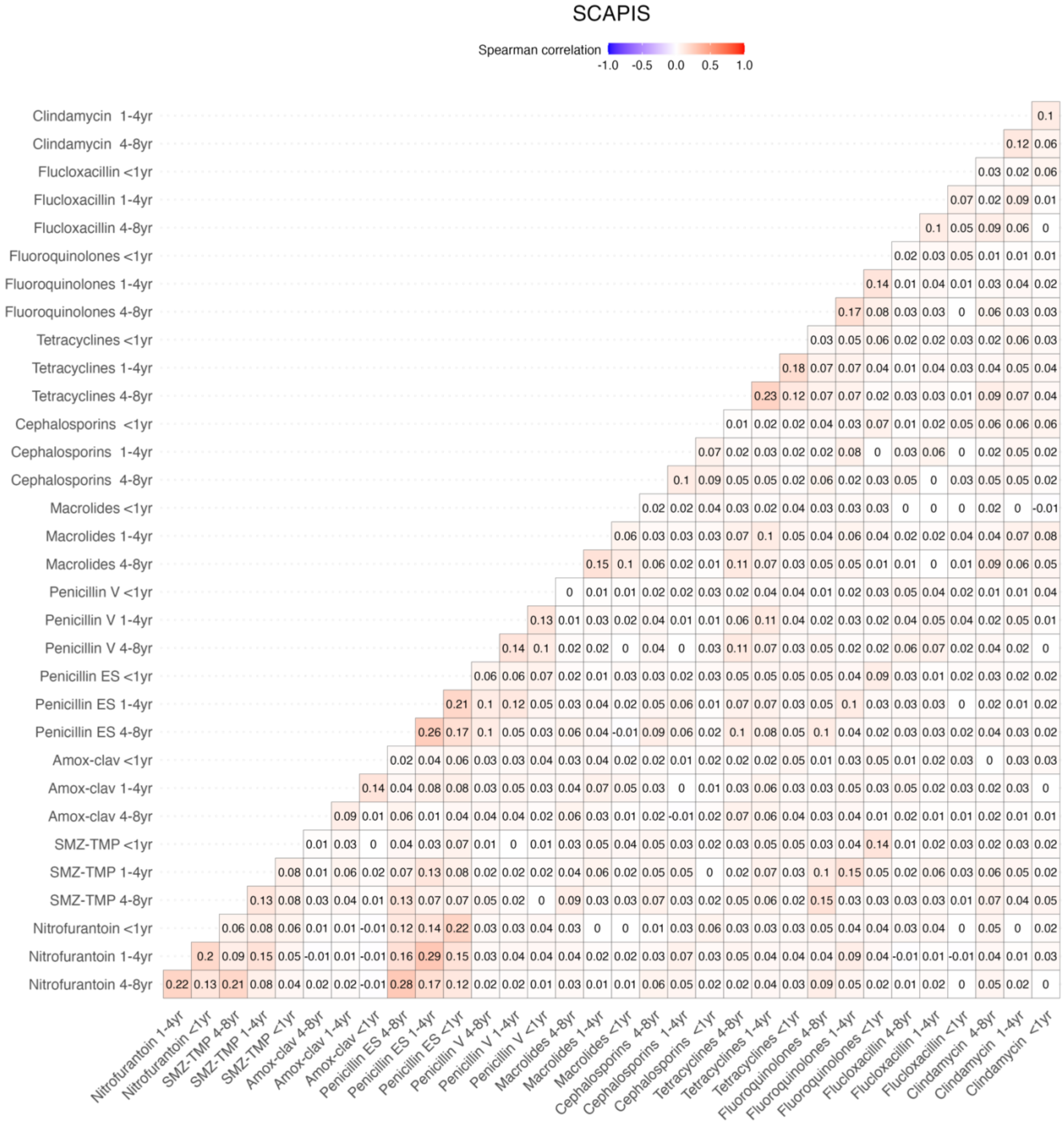

**Figure S4. Spearman Correlation in SIMPLER Between Antibiotic Classes**

Spearman correlation in SIMPLER between the number of prescriptions of each antibiotic class in the periods 4–8 years, 1–4 years, <1 year before the fecal sampling. Penicillin ES: extended-spectrum penicillins; Amox-clav: amoxicillin/clavulanic acid; SMZ-TMP: sulfamethoxazole-trimethoprim.

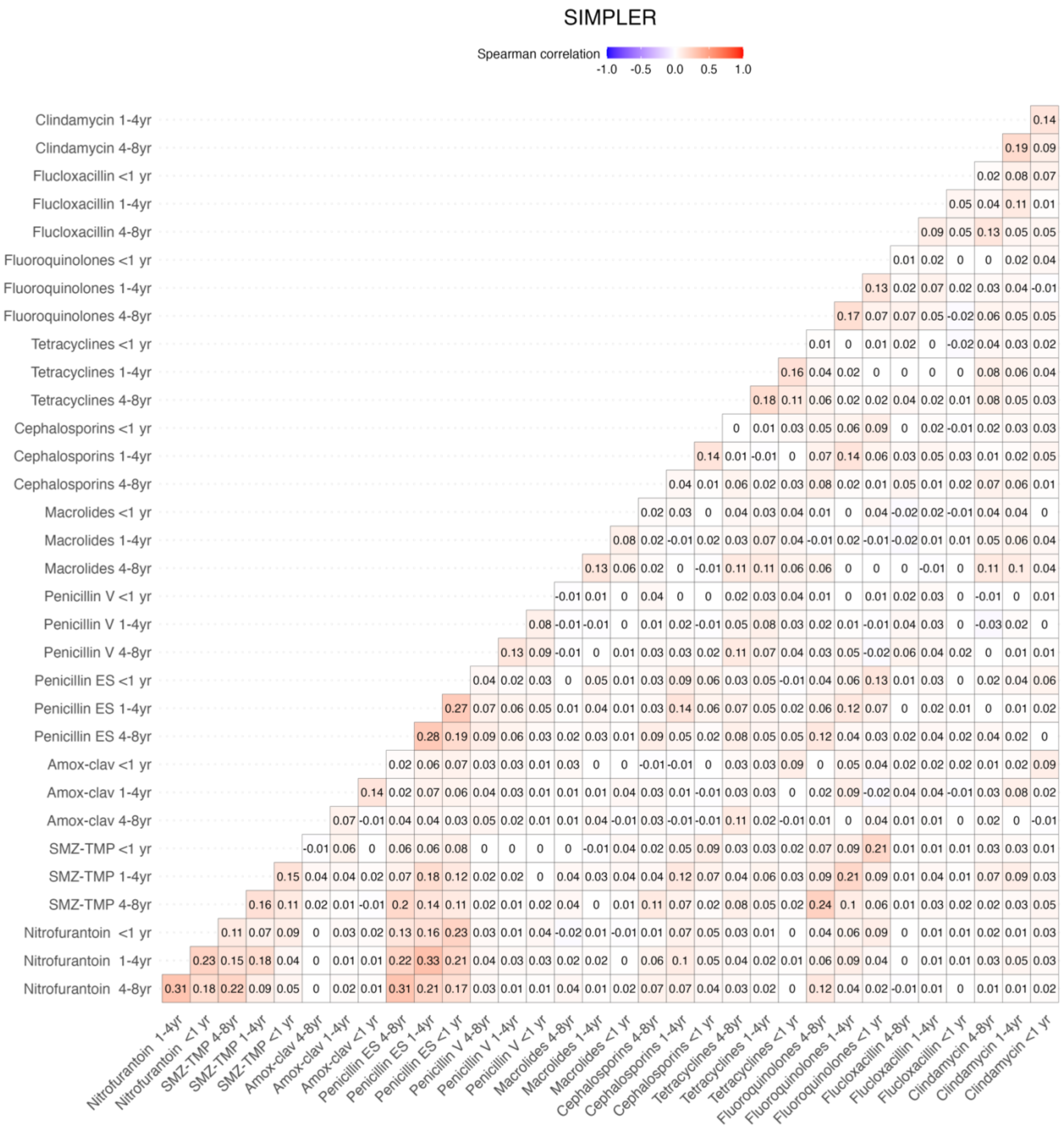

**Figure S5. Spearman Correlation in MOS Between Antibiotic Classes**

Spearman correlation in MOS between the number of prescriptions of each antibiotic class in the periods 4–8 years, 1–4 years, <1 year before the study visit. Penicillin ES: extended-spectrum penicillins; Amox-clav: amoxicillin/clavulanic acid; SMZ-TMP: sulfamethoxazole-trimethoprim**.**

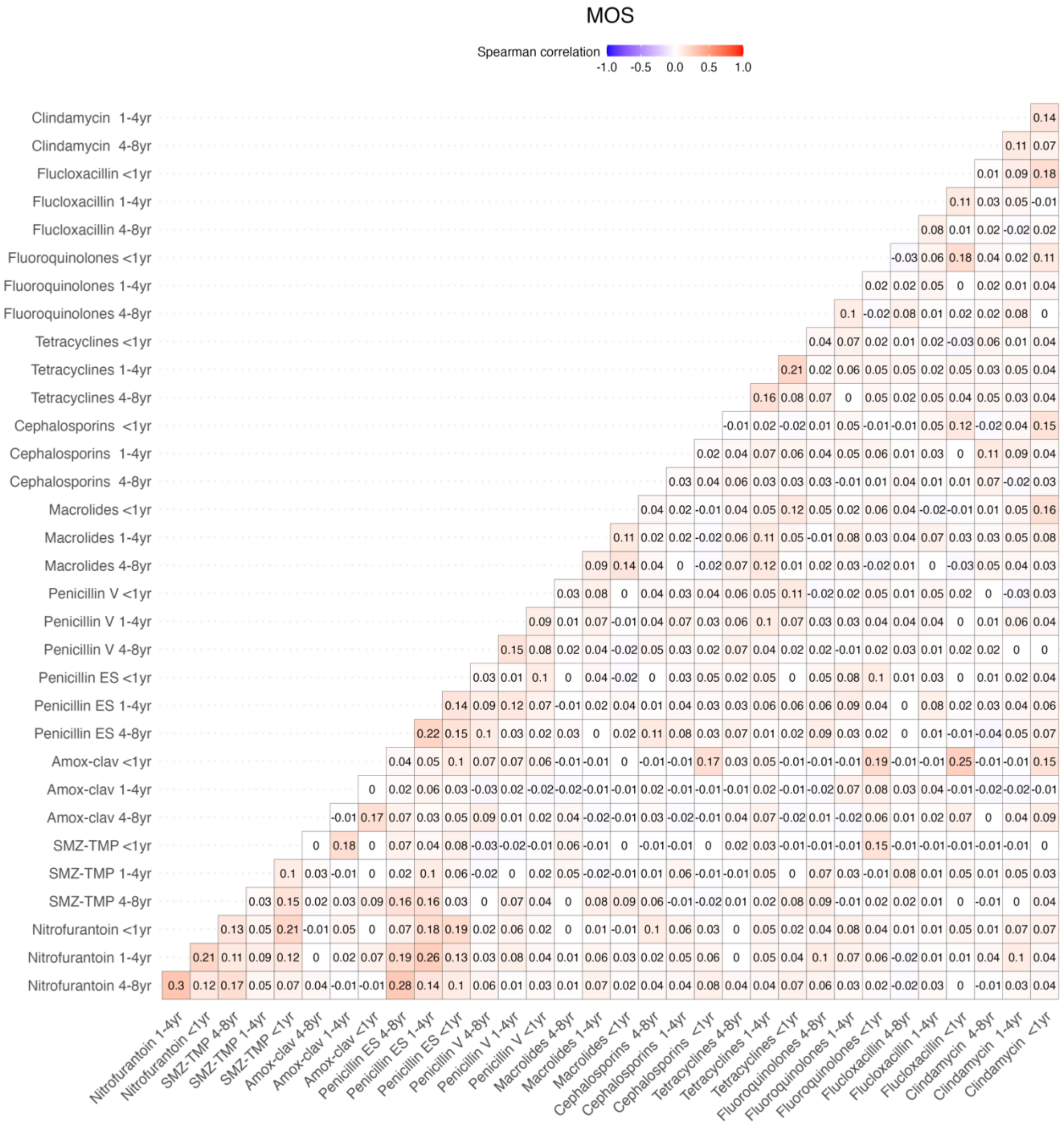

**Figure S6. Antibiotic Use and Gut Microbiota Species Diversity in Each Study and Meta-analyzed**.

Associations between antibiotic use in the 8 years before fecal sampling and gut microbiota species diversity were assessed as Shannon index, richness, and inverse Simpson index. Associations were investigated using linear regression adjusted for age, sex, smoking, education, country of birth, site-specific analysis plate, body mass index, diabetes, previous cancer diagnosis, autoimmune rheumatologic diseases, and proton-pump inhibitor use. In MOS, linear mixed models were used with family as a random effect. Meta-analyses performed using fixed-effects models. Filled shapes indicate significant associations (5% false discovery rate). Antibiotics with at least one such association are shown.

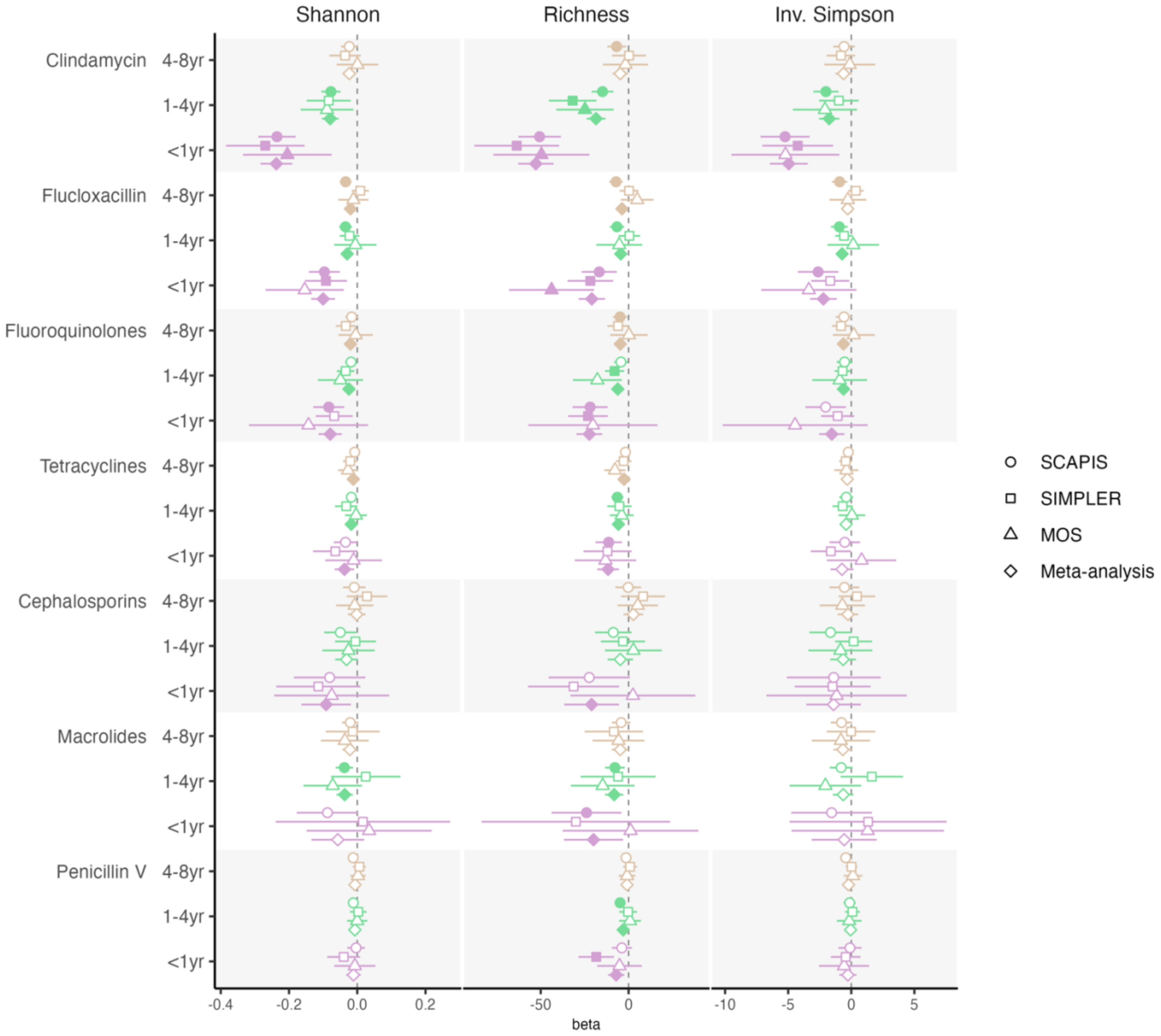

**Figure S7. Negative Control Exposures.**

Associations between antibiotic use in the 1 year after fecal sampling and gut microbiota species diversity assessed (Shannon index, richness, and inverse Simpson index). A. Associations were investigated using linear regression adjusted for antibiotic use in periods 4–8 years, 1–4 years, <1 year before fecal sampling, age, sex, smoking, education, country of birth, site-specific analysis plate, body mass index, diabetes, previous cancer diagnosis, autoimmune rheumatologic diseases, and proton-pump inhibitor use. In MOS, linear mixed models were used with family as a random effect. The x-axis represents the beta regression coefficients. Analyses were performed separately by study and meta-analyzed using fixed-effects models. B. Associations after excluding all participants with an antibiotic prescription before fecal sampling. None of the associations were significant after multiple testing adjustment (unfilled shapes). Amox-clav: Amoxicillin/Clavulanic acid. Penicillin ES: extended-spectrum penicillins; SMZ-TMP: sulfamethoxazole-trimethoprim.

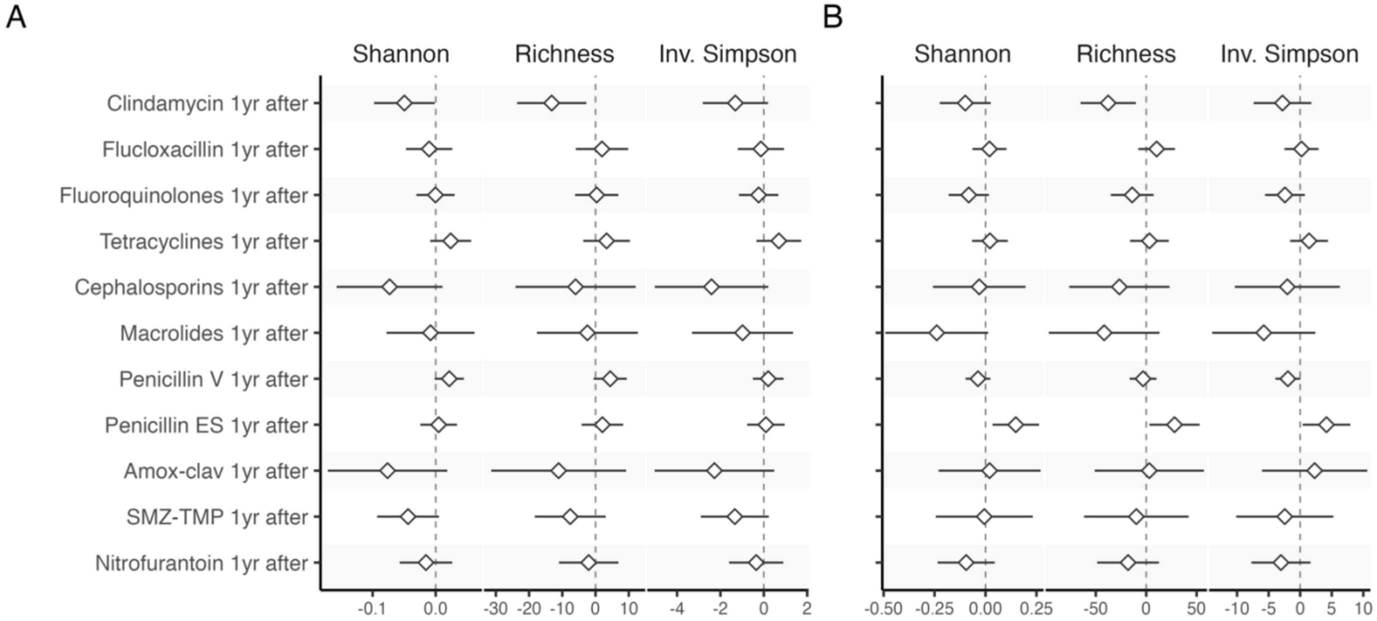

**Figure S8. Sensitivity Analyses on the Associations with Gut Microbiota Species Diversity.**

Sensitivity analysis on the associations between antibiotic use and gut microbiota species diversity assessed with Shannon index, richness, and inverse Simpson. For each cohort, we used regression models with the number of prescriptions for each antibiotic class in <1 year, 1–4 years, and 4–8 years before the fecal sampling as main exposures. Two sensitivity analyses were conducted in SCAPIS and SIMPLER: 1) removal of participants (N=550) hospitalized in the last 8 years due to infection (Table S1), and 2) removal of participants (N=5,147) hospitalized for any cause. Multiple testing was accounted for using the Benjamini-Hochberg method and reported as q-values. Full model adjusted for age, sex, smoking, education, country of birth, site-specific analysis plate, body mass index, diabetes, previous cancer diagnosis, autoimmune rheumatologic diseases, and proton-pump inhibitor use. The x-axis represents the beta regression coefficients. Filled shapes indicate significant associations (5% false-discovery rate).

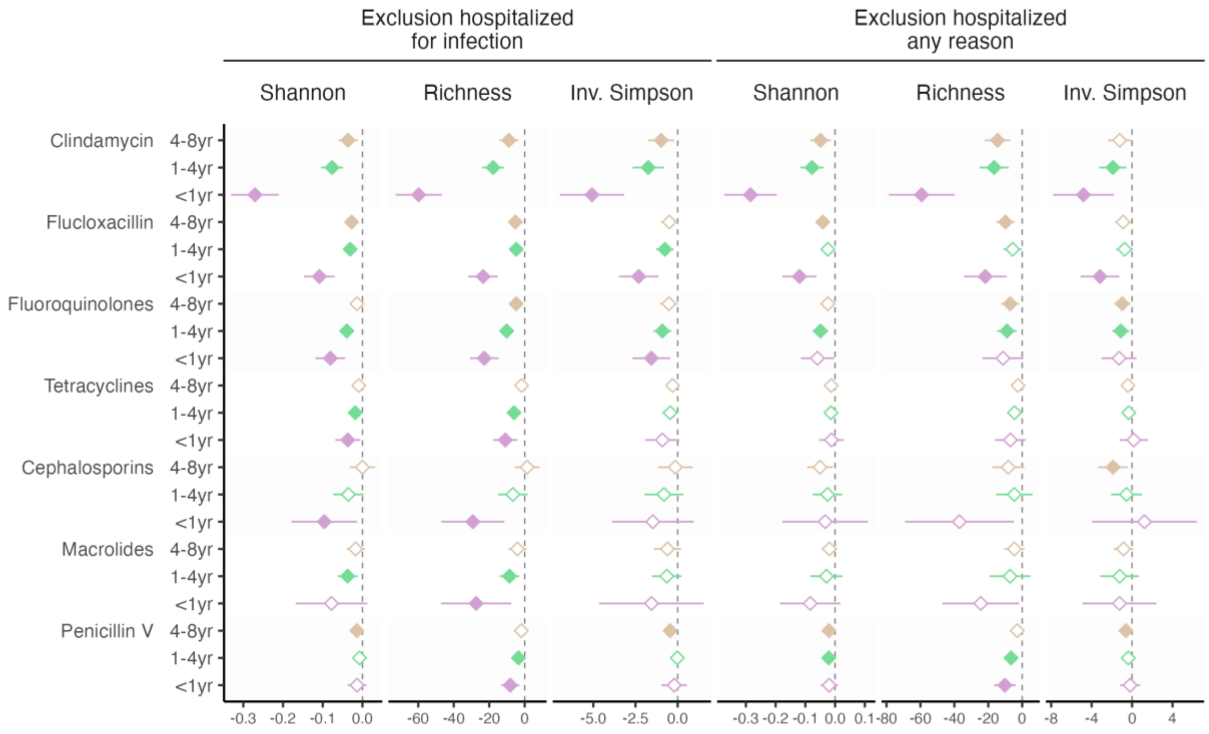

**Figure S9. A Single Antibiotic Course and Gut Microbiota Species Diversity**.

Associations between a single antibiotic course in the 8 years before fecal sampling and gut microbiota species diversity assessed (Shannon index, richness, and inverse Simpson index). Associations were investigated using linear regression adjusted for age, sex, smoking, education, country of birth, site-specific analysis plate, body mass index, diabetes, previous cancer diagnosis, autoimmune rheumatologic diseases, and proton-pump inhibitor use. In MOS, linear mixed models were used with family as a random effect. Analyses were performed separately by study and meta-analyzed using fixed-effects models. Filled shapes indicate significant associations (5% false-discovery rate). Antibiotics with at least one such association are shown. The x-axis represents the beta regression coefficients. SMZ-TMP: sulfamethoxazole-trimethoprim.

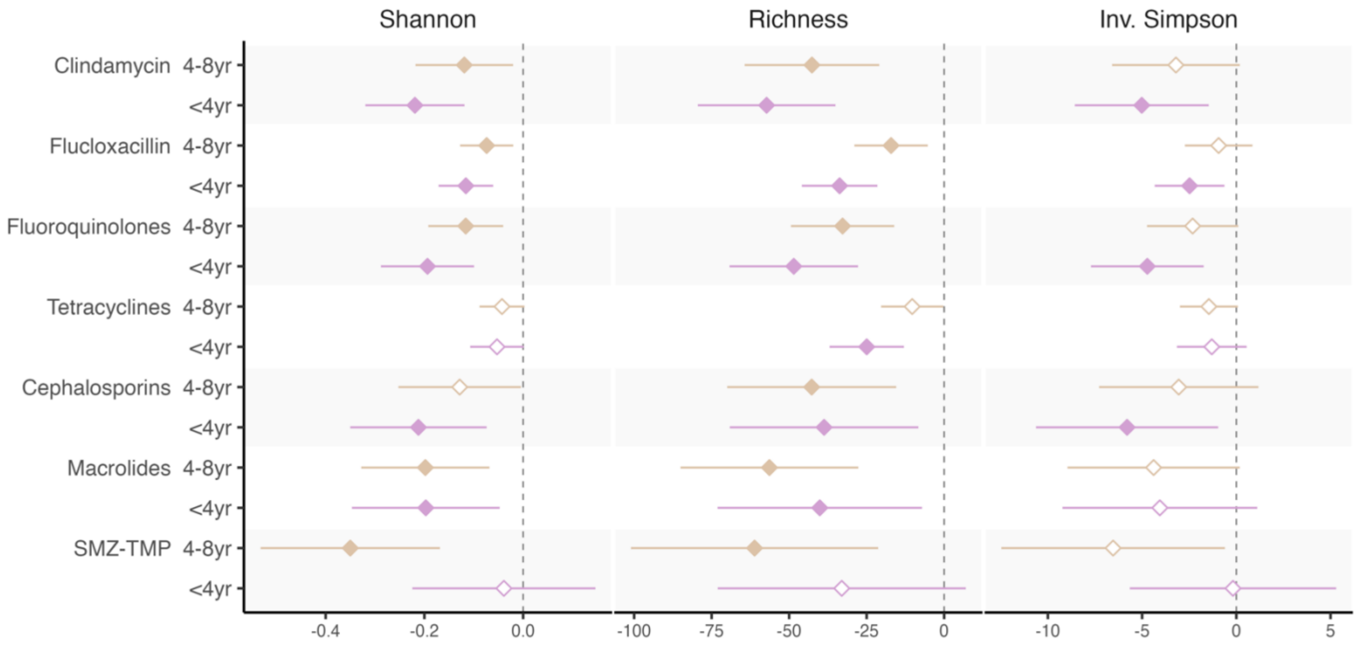

**Figure S10. Sensitivity Analysis on the Associations with Gut Microbiota Species Abundance**

Associations between antibiotic use in the 8 years before fecal sampling and gut microbiota species abundances were investigated (A) after removing 550 individuals who had been hospitalized due to an infection (Table S1) in the last 8 years in SCAPIS and SIMPLER, and (B) after removing 5,147 individuals who had been hospitalized due to any reason in the last 8 years in SCAPIS and SIMPLER. The number of prescriptions in the three periods (4–8 years, 1–4 years, <1 year) and for each antibiotic were included in the same regression model. Analyses were performed in each study and meta-analyzed using fixed-effects models. Only significant associations at a 5% false discovery rate are shown. Red circles indicate positive associations (increased species abundance), and blue circles indicate negative associations (decreased species abundance). Full model adjusted for age, sex, smoking, education, country of birth, site-specific analysis plate, body mass index, diabetes, previous cancer diagnosis, autoimmune rheumatologic diseases, and proton-pump inhibitor use. The numbers above the circles indicate individuals who used the antibiotic at least once in that period. Amox-clav: Amoxicillin/Clavulanic acid, Penicillin ES: extended-spectrum penicillins; SMZ-TMP: sulfamethoxazole-trimethoprim.

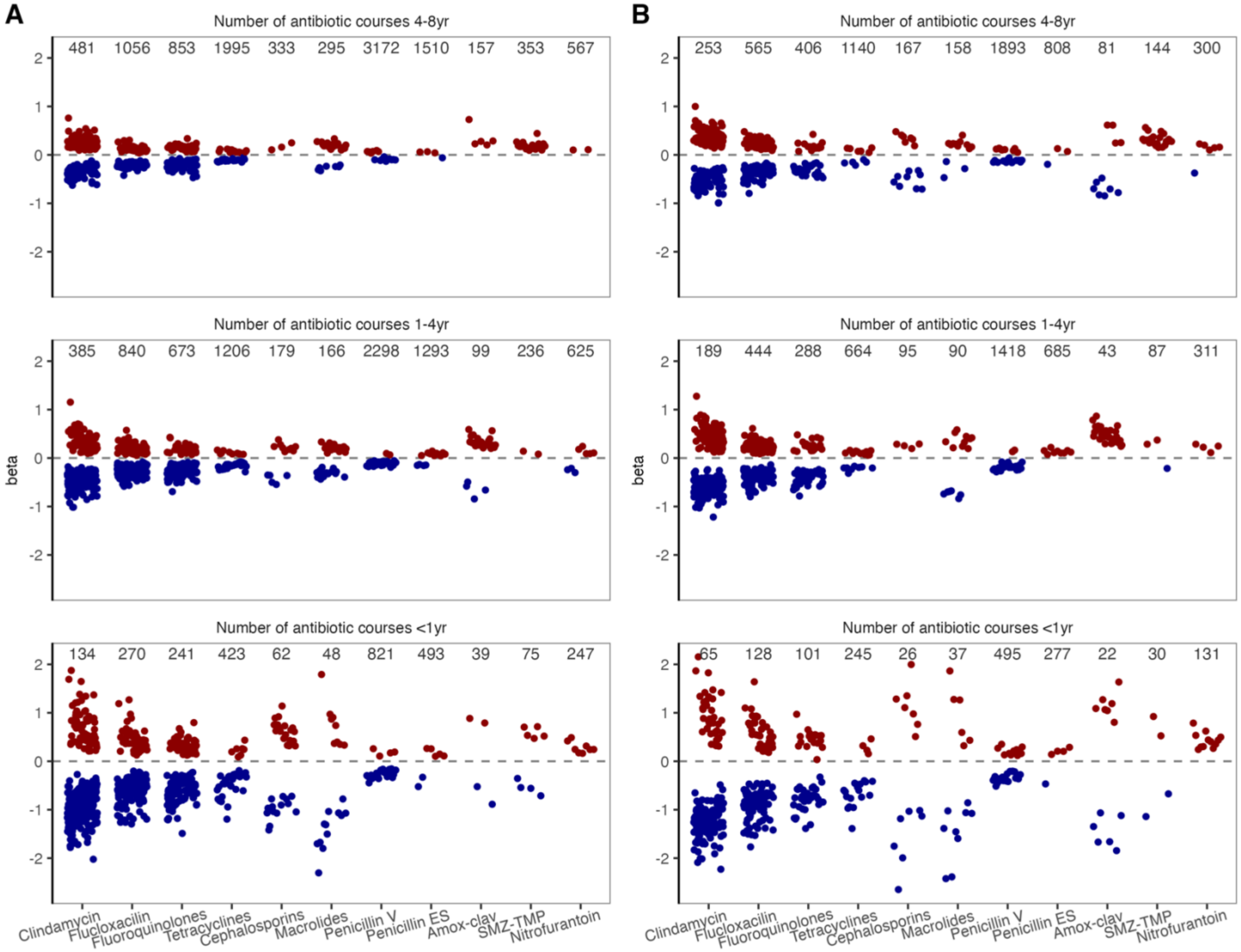

329
